# SARS-CoV-2 infection in primary schools in northern France: A retrospective cohort study in an area of high transmission

**DOI:** 10.1101/2020.06.25.20140178

**Authors:** Arnaud Fontanet, Rebecca Grant, Laura Tondeur, Yoann Madec, Ludivine Grzelak, Isabelle Cailleau, Marie-Noëlle Ungeheuer, Charlotte Renaudat, Sandrine Fernandes Pellerin, Lucie Kuhmel, Isabelle Staropoli, François Anna, Pierre Charneau, Caroline Demeret, Timothée Bruel, Olivier Schwartz, Bruno Hoen

## Abstract

**Background:** The extent of SARS-CoV-2 transmission among pupils in primary schools and their families is unknown.

**Methods:** Between 28-30 April 2020, a retrospective cohort study was conducted among pupils, their parents and relatives, and staff of primary schools exposed to SARS-CoV-2 in February and March 2020 in a city north of Paris, France. Participants completed a questionnaire that covered sociodemographic information and history of recent symptoms. A blood sample was tested for the presence of anti-SARS-CoV-2 antibodies using a flow-cytometry-based assay.

**Results:** The infection attack rate (IAR) was 45/510 (8.8%), 3/42 (7.1%), 1/28 (3.6%), 76/641 (11.9%) and 14/119 (11.8%) among primary school pupils, teachers, non-teaching staff, parents, and relatives, respectively (P = 0.29). Prior to school closure on February 14, three SARS-CoV-2 infected pupils attended three separate schools with no secondary cases in the following 14 days among pupils, teachers and non-teaching staff of the same schools. Familial clustering of cases was documented by the high proportion of antibodies among parents and relatives of infected pupils (36/59 = 61.0% and 4/9 = 44.4%, respectively). In children, disease manifestations were mild, and 24/58 (41.4%) of infected children were asymptomatic.

**Interpretation:** In young children, SARS-CoV-2 infection was largely mild or asymptomatic and there was no evidence of onwards transmission from children in the school setting.

## Introduction

As the coronavirus (COVID-19) pandemic continues to evolve, the extent of SARS-CoV-2 infection in children has not been well documented and the role children may play in virus transmission remains unclear. During the first epidemic wave, many countries included school closures among the measures implemented to limit viral transmission, largely based on the evidence of the impact of school closures on influenza transmission^1,2^. As many schools are now reopening, it is critical to evaluate the risk of viral circulation among pupils and their teachers in schools^3^.

Initial epidemiological data from China indicated that children were significantly less affected than adults, whether considering the total number of clinical cases, disease severity or fatal outcomes^4^. Similar findings were also reported in other countries^5-7^. It is now understood that children, when infected, present with mild and asymptomatic forms of the disease more often than adults^8-10^, with severe and fatal outcomes remaining rare in children^11-12^.

Younger children are generally thought to be less susceptible to infection as compared to adults^13-17^, and, when infected, are usually contaminated by a household member^18^. Some studies have nevertheless documented secondary attack rates in families as high in children as in adults^19^. Children, when infected, may carry the virus in their throats for 9-11 days^18^ and for up to one month in stools^20^. Viral loads have been found to be similar between infected children and adults^21-22^, which would suggest that children may be as infectious as adults. It is therefore unclear why children would be less susceptible, and less infectious, as compared to adults^23^. Seroepidemiological studies are thus needed to determine the extent of infection in children and to decipher the role they may play in transmission

To our knowledge, the number of SARS-CoV-2 secondary transmissions in school setting documented in scientific literature is limited, with very few or no secondary cases in investigations in Australia^24^, Ireland^25^, and France^26^, with the exception of one important cluster in a high school north of Paris in February 2020^27^.

It is all the more important to understand the extent of infection in children and the role they may play in transmission given the likely negative effects of school closures on educational achievement and economic outcomes^3^. To investigate the extent of infection in younger children, a follow-up seroepidemiologic investigation to that in the high school in a city north of Paris was conducted across primary schools in the same city. Here, we present the results of the follow up investigation in primary schools.

## Methods

### Study setting

An initial retrospective epidemiological investigation was conducted in the Crépy-en-Valois city (15,000 inhabitants) north of Paris, France after the diagnosis of the first case of COVID-19 on 24 February 2020. This investigation identified an epidemic around a local high school with two teachers having symptoms consistent with COVID-19 as early as on 2 February 2020. Since there was no known circulation of SARS-CoV-2 at that time in the region, no public health or social measures intended to limit the viral transmission had been implemented and no active SARS-CoV-2 testing had been conducted. A preliminary rapid investigation among symptomatic adults and pupils at the high school on 5-6 March 2020 revealed that 11/66 (16.7%) adults and 2/24 (8.3%) pupils had acute infection, as determined by a positive RT-PCR test result. As a follow-up to this rapid investigation, the decision was made to further examine by serological testing the extent of infection among pupils, their parents and relatives, teaching staff and non-teaching staff of 1) the high school where the two teachers worked and 2) the primary schools in the same city. The high school investigation has previously been reported^26^. Here, we describe the follow-up seroepidemiologic investigation across six primary schools from the same city, among children aged 6 to 11 years.

### Study design

A retrospective cohort study was conducted by inviting all pupils, teachers and non-teaching staff (administrative, cleaners, catering) from each of the six primary schools who were registered at the school from the beginning of the epidemic (estimated around 13 January 2020) up to the time of the investigation (28-30 April 2020). Since pupils were minor, at least one parent was invited to provide informed consent for their child. The parents were also invited to participate in the study, as well as any of the other children or relatives over the age of 5 years living in the same household.

Following informed consent, participants (with the help of their parents in the case of pupils) completed a questionnaire that covered sociodemographic information, underlying medical conditions, history of recent symptoms, and history of COVID-19 diagnosis prior to this investigation. A 5-mL blood sample was taken from all participants.

### Laboratory analyses

Samples were transferred to and stored in the Clinical Investigation and Access to BioResources (ICAReB) biobank platform at the Institut Pasteur (Paris, France), which collects and manages human bioresources for scientific purposes, following ISO 9001 and NF S 96-900 quality standards (BRIF code n°BB-0033-00062). Serological testing was conducted using the S-Flow assay, a flow-cytometry-based serological test developed by the Institut Pasteur. The assay is based on the recognition of the SARS-CoV-2 Spike protein expressed at the surface of 293T cells. In previous studies, the sensitivity of the assay was estimated at 99.4% (95% confidence interval (CI) = 96.6% - 100%) on a panel of 160 RT-PCR confirmed mild forms of COVID-19^28^, while its specificity was found to be 100% (one-sided 97.5% CI = 97.4% - 100%) on a panel of 140 pre-epidemic sera^29^.

### Case definitions

Any participant with a positive serology at the time of blood sampling was considered as having had SARS-CoV-2 infection. Each infection was categorized as symptomatic if any recent symptoms were reported by the participant up until 7 days prior to the date of sample collection to allow sufficient time for seroconversion^30,31^, or, alternatively, as asymptomatic. Symptoms were further categorized as major (fever, dry cough, dyspnea, anosmia and ageusia) or minor (sore throat, rhinitis, muscle pain, diarrhea, headache, asthenia, vomiting, nausea, chest pain, abdominal pain).

### Statistical analyses

The IAR was defined as the proportion of all participants with SARS-CoV-2 infection based on antibody detection in the collected blood sample by the end of the first COVID-19 epidemic wave. Participants were further categorized as children if under 18 years of age (pupils, relatives of the pupil living in the same household) and adults if 18 years or older. The analysis was also broken down by school, and by time period (before and after February 14, date of the school closure for two-week holidays immediately followed by a local lockdown on 1 March 2020). The IAR was compared according to participants’ characteristics using chi-square test and Fisher exact test, where appropriate. All statistical analyses were performed using Stata 15.0 (StataCorp, College Station, TX, USA).

### Ethical considerations

This study is included as part of the registration on ClinicalTrials.gov under the identifier NCT04325646, cohort ‘CORSER-2c,’ and received ethical approval by the Comité de Protection des Personnes Ile de France III. Informed consent was obtained from all participants.

## Results

From 28 to 30 April 2020, 1047 pupils and 51teachers, from six primary schools, with children aged 6 to 11 years, were invited by email to participate in the investigation. Of these, 541 (51.5%) pupils and 46 (90.2%) teachers accepted to participate in the study. Thirty-one pupils were excluded as they refused phlebotomy, as were four teachers not directly affiliated with any of the six schools. This resulted in 510 pupils and 42 teachers with blood sample to be analyzed. In addition, 641 parents of pupils, 119 relatives of pupils sharing the same household, and 28 non-teaching staff completed the study population (Figure 1). Table 1 indicates the characteristics of the 1340 participants. Pupils and their parents constituted the majority of the study population (38.1% and 47.8%, respectively).

**Table 1.**
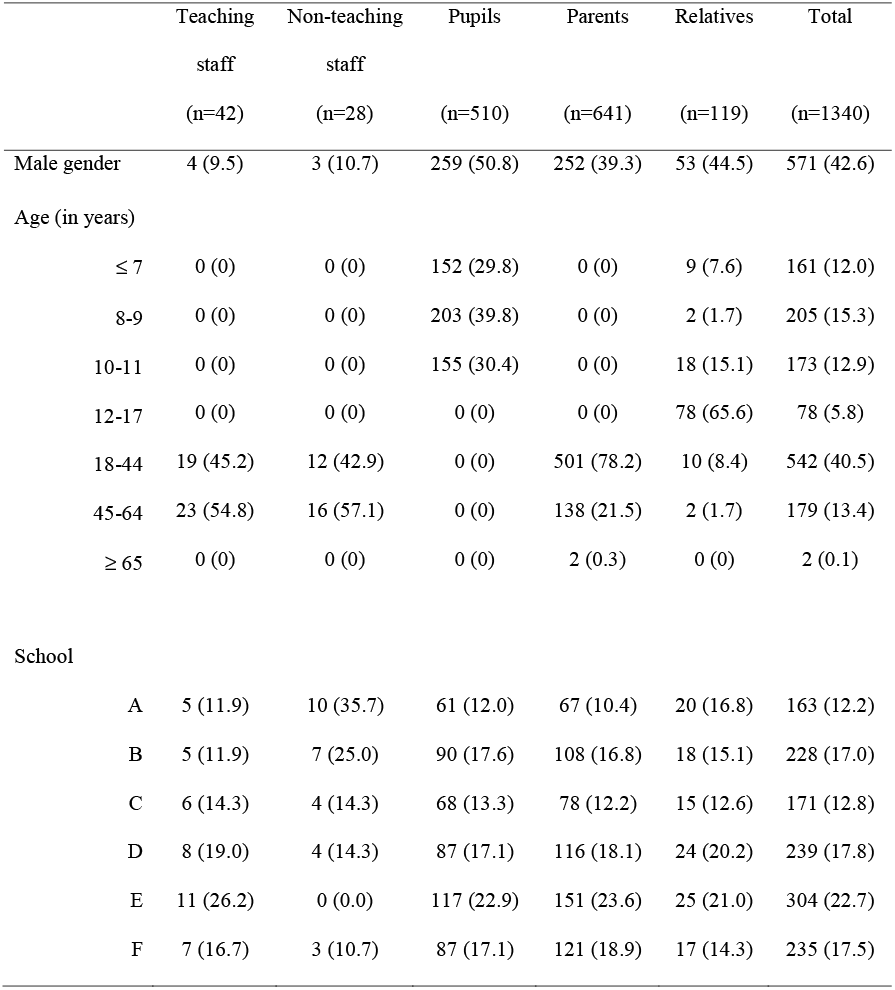
Sociodemographic characteristics of the 1340 participants of the SARS-CoV-2, France from 28-30 April 2020

|  | Teaching<br>staff<br>(n=42) | Non-teaching<br>staff<br>(n=28) | Pupils<br>(n=510) | Parents<br>(n=641) | Relatives<br>(n=119) | Total<br>(n=1340) |
| --- | --- | --- | --- | --- | --- | --- |
| Male gender | 4 (9.5) | 3 (10.7) | 259 (50.8) | 252 (39.3) | 53 (44.5) | 571 (42.6) |
| Age (in years) |  |  |  |  |  |  |
| ≤ 7 | 0 (0) | 0 (0) | 152 (29.8) | 0 (0) | 9 (7.6) | 161 (12.0) |
| 8-9 | 0 (0) | 0 (0) | 203 (39.8) | 0 (0) | 2 (1.7) | 205 (15.3) |
| 10-11 | 0 (0) | 0 (0) | 155 (30.4) | 0 (0) | 18 (15.1) | 173 (12.9) |
| 12-17 | 0 (0) | 0 (0) | 0 (0) | 0 (0) | 78 (65.6) | 78 (5.8) |
| 18-44 | 19 (45.2) | 12 (42.9) | 0 (0) | 501 (78.2) | 10 (8.4) | 542 (40.5) |
| 45-64 | 23 (54.8) | 16 (57.1) | 0 (0) | 138 (21.5) | 2 (1.7) | 179 (13.4) |
| ≥ 65 | 0 (0) | 0 (0) | 0 (0) | 2 (0.3) | 0 (0) | 2 (0.1) |
| School |  |  |  |  |  |  |
| A | 5 (11.9) | 10 (35.7) | 61 (12.0) | 67 (10.4) | 20 (16.8) | 163 (12.2) |
| B | 5 (11.9) | 7 (25.0) | 90 (17.6) | 108 (16.8) | 18 (15.1) | 228 (17.0) |
| C | 6 (14.3) | 4 (14.3) | 68 (13.3) | 78 (12.2) | 15 (12.6) | 171 (12.8) |
| D | 8 (19.0) | 4 (14.3) | 87 (17.1) | 116 (18.1) | 24 (20.2) | 239 (17.8) |
| E | 11 (26.2) | 0 (0.0) | 117 (22.9) | 151 (23.6) | 25 (21.0) | 304 (22.7) |
| F | 7 (16.7) | 3 (10.7) | 87 (17.1) | 121 (18.9) | 17 (14.3) | 235 (17.5) |

**Figure 1.**
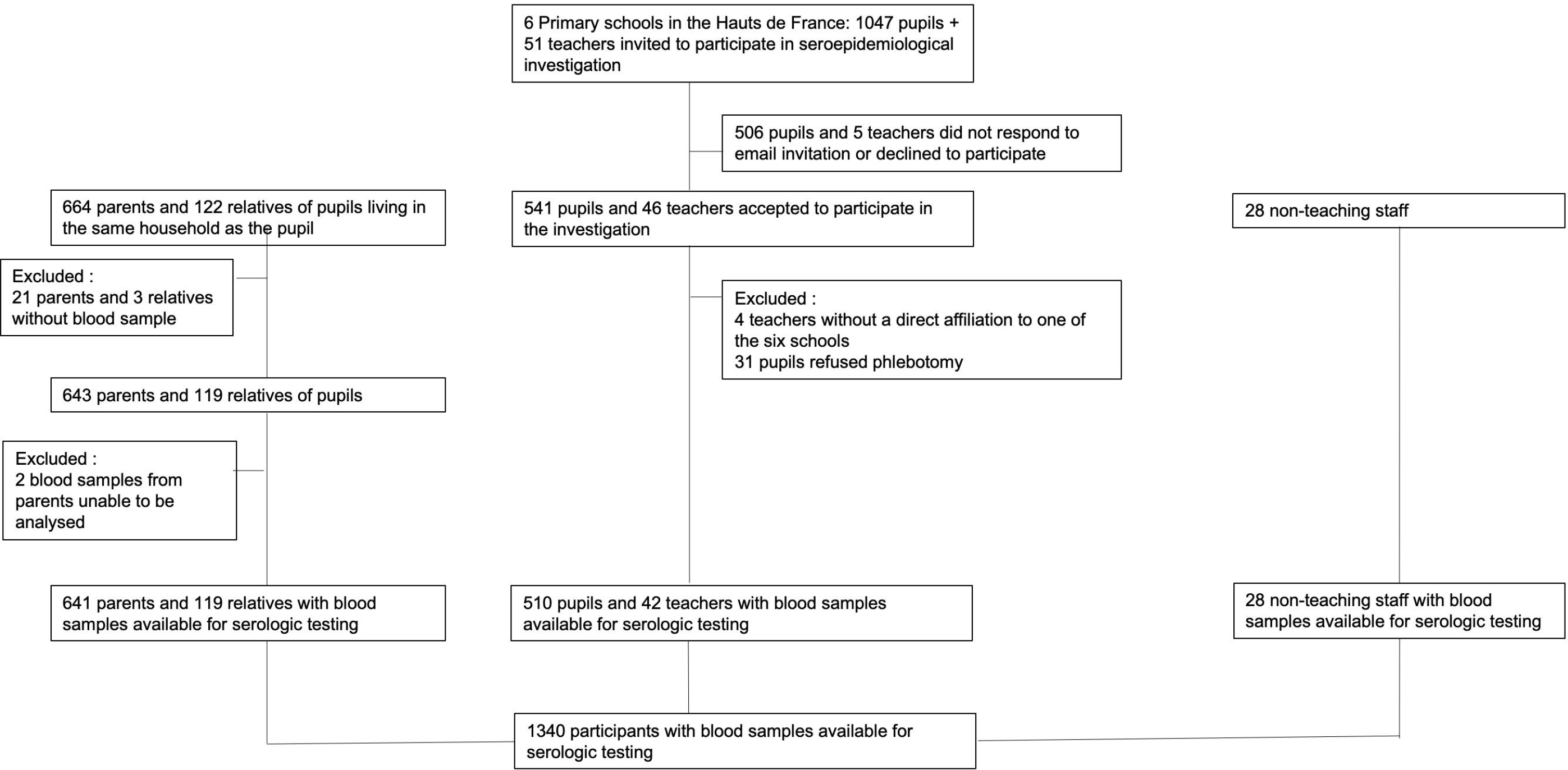
Flowchart of enrolment of participants

Most participants were female (57.4%), particularly among teaching (90.5%) and non-teaching (89.3%) staff. The pupils were aged 6-11 years, while the median (IQR) age was 40 (37-44) years for parents, 47.5 (40-51) years for teachers, and 47.5 (32-54) years for non-teaching staff (Table 1).

The overall IAR across study participants was 139/1340 (10.4%). It did not differ by gender, age categories, or type of participants (Table 2). The epidemic curve, based on symptoms experienced by participants with SARS-CoV-2 antibodies, had no specific pattern, and transmission does not appear to have been impacted by the closure of schools for holidays on February 14 (end of week 7) (Figure 2A). There were three instances in three separate schools of high suspicion of SARS-CoV-2 infection in pupils before the closure of the school for holidays (end of week 7), one in week 6, and two in week 7. There were no secondary cases in pupils, teachers and non-teaching staff of the corresponding schools in the 14 days following these initial cases, except for one teacher who had onset of symptoms nine days later, but also had a close contact with a confirmed case outside of the school five days prior to becoming sick. Parents of infected pupils had higher IAR compared to parents of non-infected pupils (61.0% versus 6.9%; P <0.0001), and relatives of infected pupils had higher IAR compared to relatives of non-infected pupils (44.4% versus 9.1%; P = 0.002) (Table 2 & Figure S1).

**Table 2.** Infection attack rate (IAR) according to sociodemographic characteristics

|  | N | n (%) | P value |
| --- | --- | --- | --- |
| Gender |  |  |  |
| Male | 571 | 54 (9.5) | 0.34 |
| Female | 769 | 85 (11.1) |  |
| Age (in years) |  |  |  |
| ≤ 7 | 161 | 10 (6.2) | 0.36 |
| 8-9 | 205 | 20 (9.8) |  |
| 10-11 | 173 | 16 (9.2) |  |
| 12-17 | 78 | 12 (15.4) |  |
| 18-44 | 542 | 62 (11.4) |  |
| 45-64 | 179 | 19 (10.6) |  |
| ≥ 65 | 2 | 0 (0.0) |  |
| Type of participant |  |  |  |
| Pupil | 510 | 45 (8.8) | 0.29 |
| Teacher | 42 | 3 (7.1) |  |
| Non-teaching staff | 28 | 1 (3.6) |  |
| All parents | 641 | 76 (11.9) |  |
| Parent of an infected pupil | 59 | 36 (61.0) |  |
| Parent of a non-infected pupil | 582 | 40 (6.9) |  |
| All relatives living in the same household | 119 | 14 (11.8) |  |
| Relatives of an infected pupil | 9 | 4 (44.4) |  |
| Relatives of a non-infected pupil | 110 | 10 (9.1) |  |

**Figure 2.**
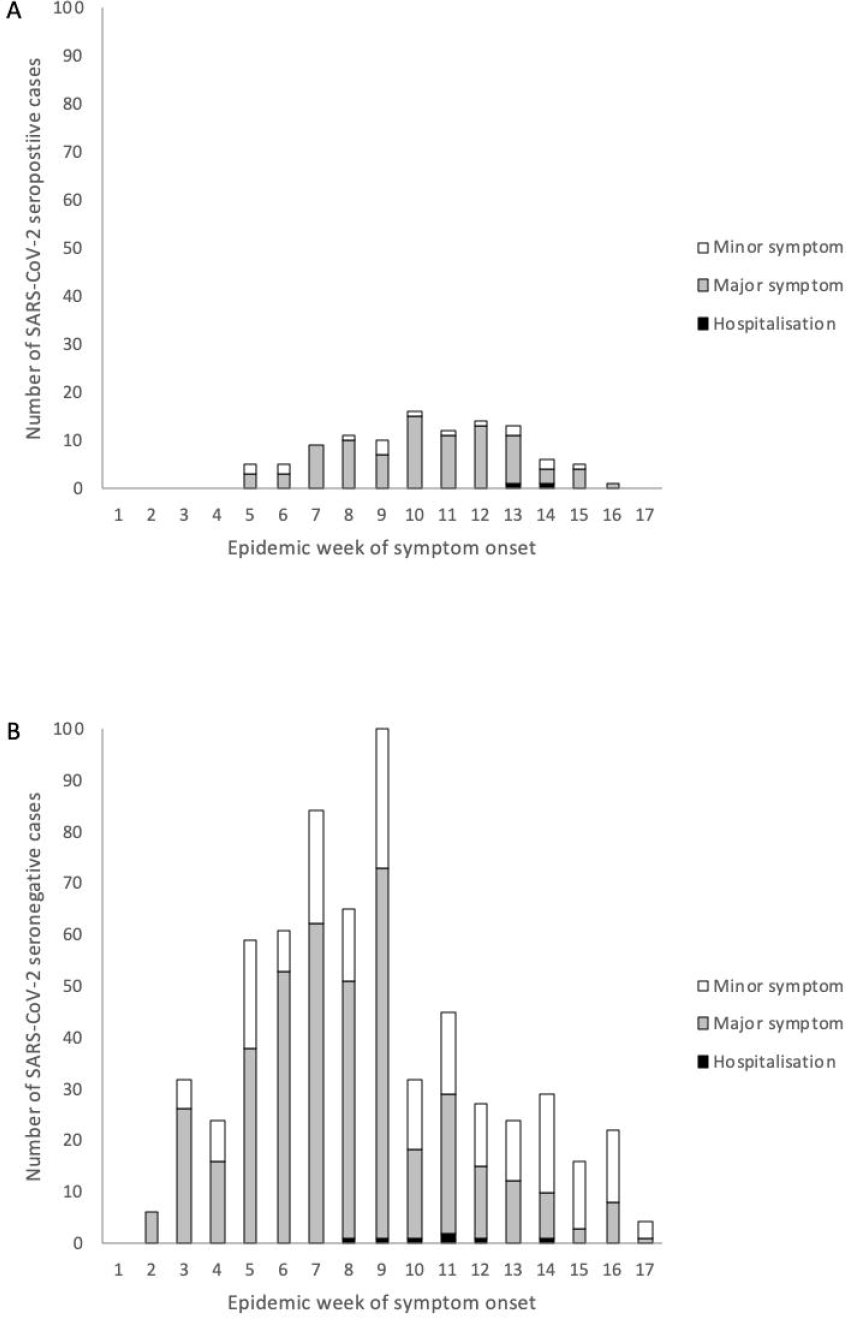
Timeline of symptom onset among (A) 107 symptomatic individuals who were seropositive for anti-SARS-CoV-2 antibodies; (B) 631 symptomatic individuals who were seronegative for anti-SARS-CoV-2 antibodies.

Among adults, fever, cough, dyspnea, ageusia, anosmia, muscle pain, sore throat, headache, asthenia, and diarrhea were all associated with the detection of SARS-CoV-2 antibodies (Table 3). Ageusia and anosmia, reported among 48% of adult participants, had a high positive predictive value for infection: 75.0% and 90.7%, respectively. In children, only asthenia and diarrhea were associated with SARS-CoV-2 antibodies. Only two children with SARS-CoV-2 antibodies experienced anosmia and ageusia, and both were 15 years of age. Among the 139 participants with SARS-CoV-2 antibodies, only two (1.4%, 95% CI = 0.2% - 5.1%), both parents, were hospitalized. There were no deaths. Across the study period, 9.9% of seropositive adults, and 41.4% of seropositive children, reported no symptoms (P <0.001). Symptoms of respiratory infections – fever, cough, rhinitis – were common among the participants without SARS-CoV-2 antibodies during the study period, with a marked decrease after lockdown was introduced on 1 March 2020 (Figure 2B).

**Table 3.**
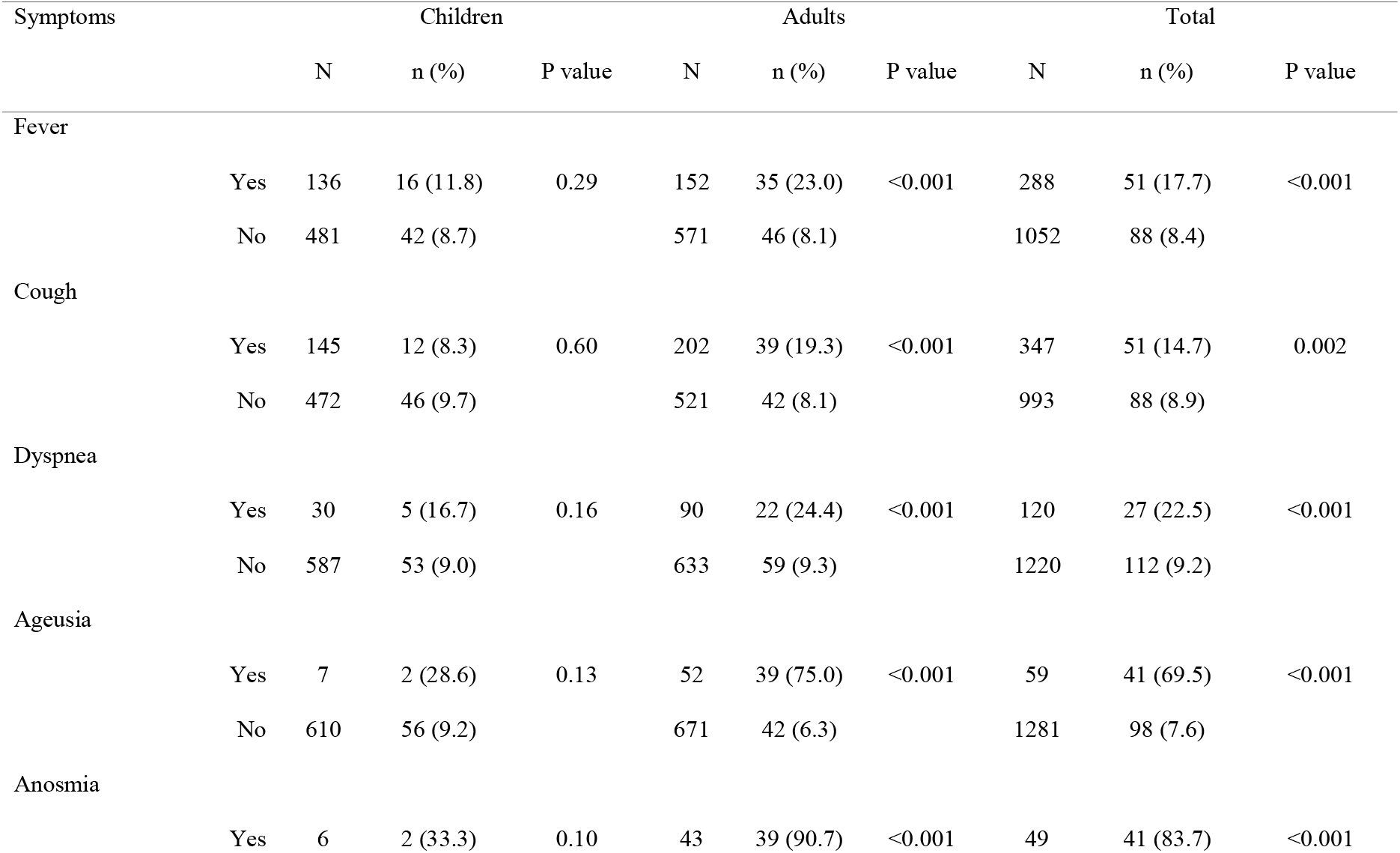
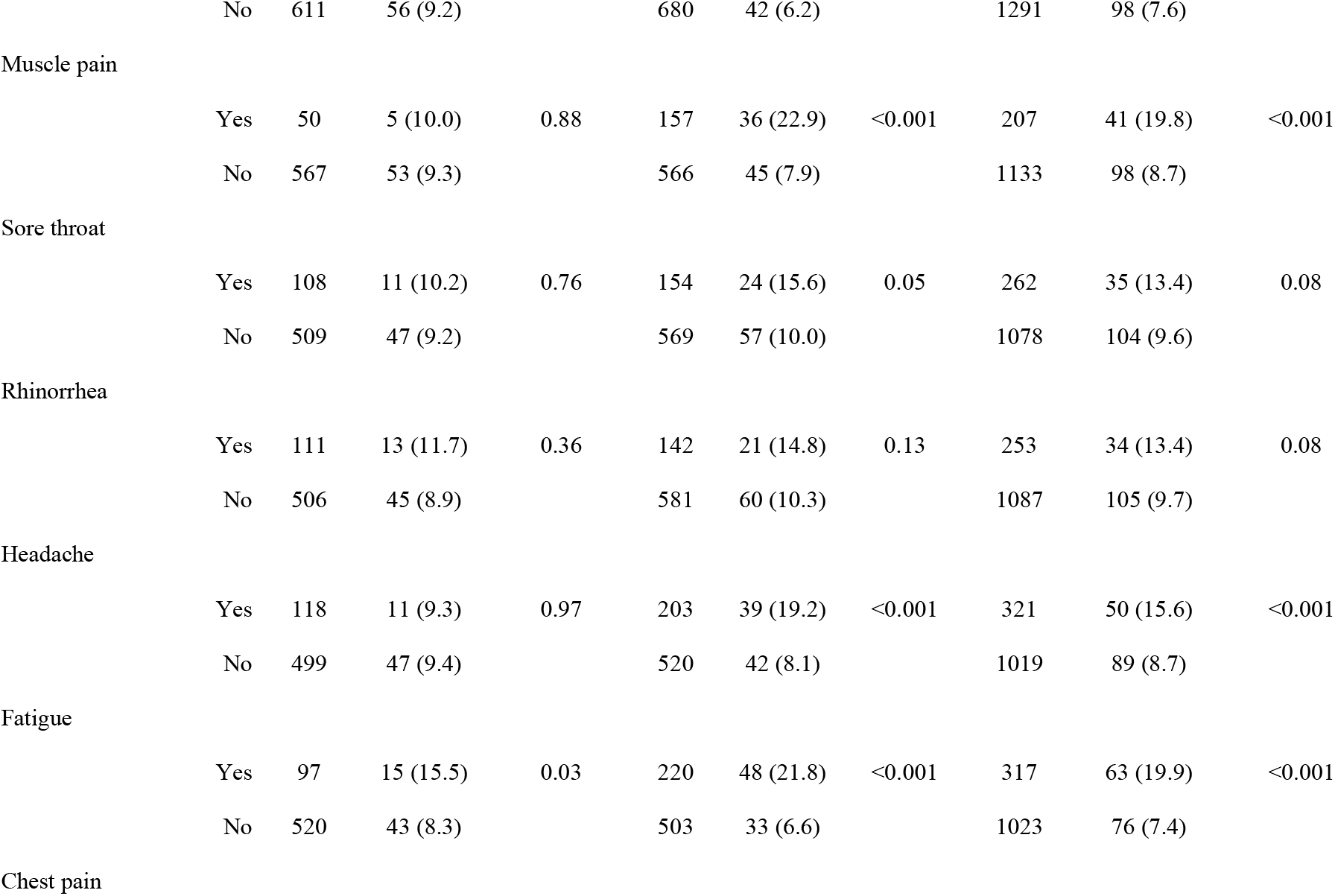
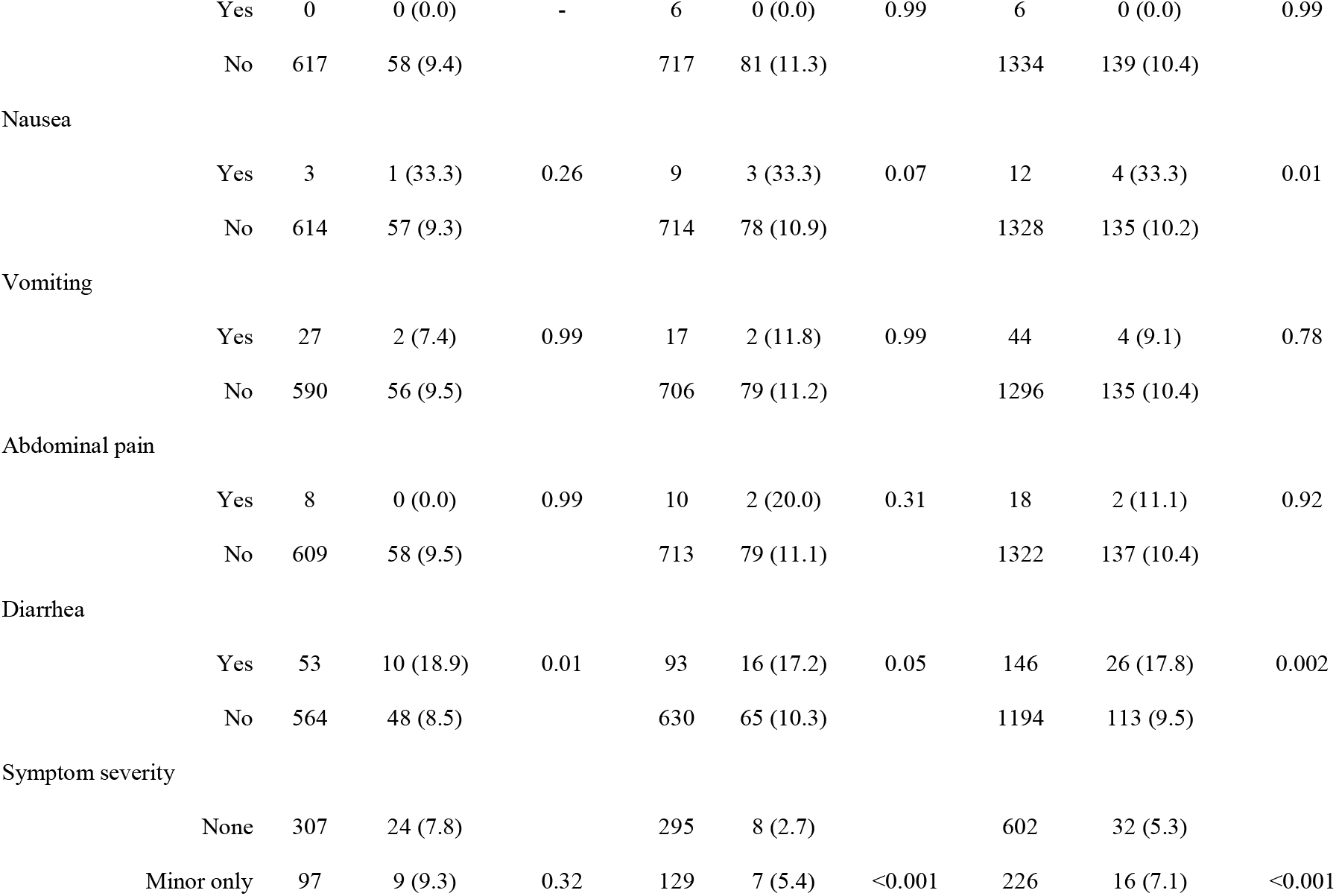
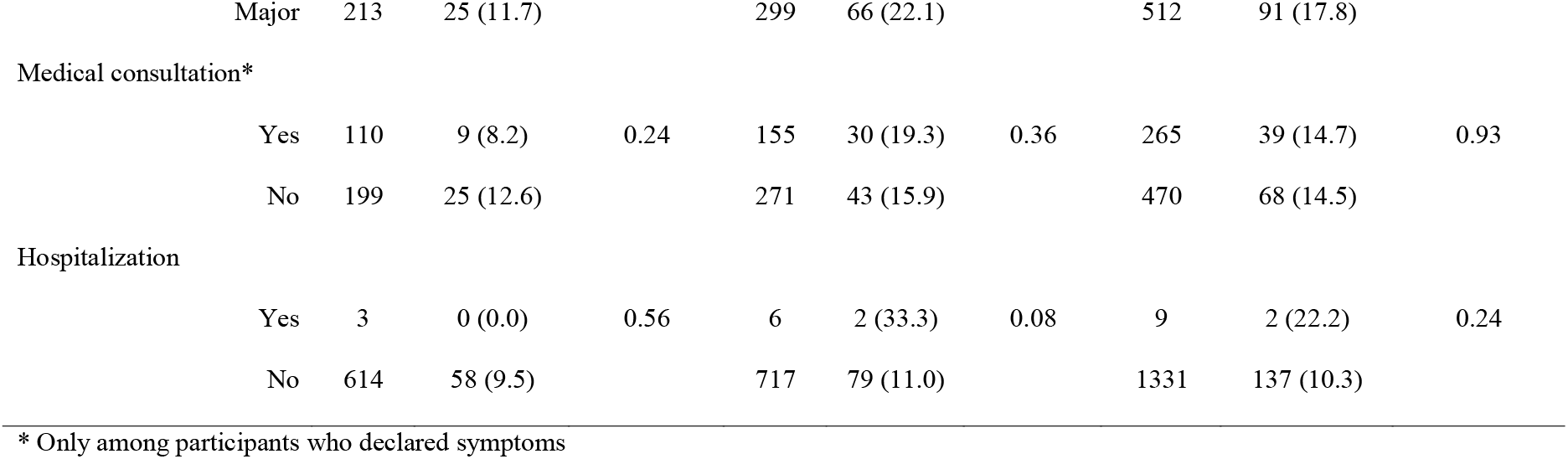
IAR (%) by symptoms and type of participant

## Discussion

This study in one of the first seroepidemiologic investigations on SARS-CoV-2 in the primary school setting. Despite three introductions of the virus into three primary schools, there appears to have been no further transmission of the virus to other pupils or teaching and non-teaching staff of the schools. In families of infected pupils, the prevalence of antibodies was very high, suggesting intrafamilial clustering of infections. Finally, children experienced mild forms of disease, with many of them being asymptomatic.

We can infer from the reported date of symptom onset among seropositive individuals that viral circulation in the study population presumably began around week 5 (27-31 January 2020). Transmission continued to increase up to week 10 (2-6 March), with no effect of school closure for holidays (14 February). Transmission stabilized and then declined after week 13 (23-27 March). Since there was no reported circulation of SARS-CoV-2 during the month of February in the region until the diagnosis of the first local case on 24 February 2020, adherence to any public health or social measures intended to limit the transmission of the virus was likely low, allowing us to document the natural circulation of the virus in the community.

We could identify three symptomatic SARS-CoV-2 infected pupils in three separate schools during the three weeks preceding closure of the school for holidays and then lockdown. There were no secondary cases in pupils, teachers and non-teaching staff of the corresponding schools in the 14 days following these initial cases. These findings are in line with previous studies from Australia^24^, Ireland^25^, and France^26^. They differ however from the results of the study performed in the high school of the same city in France, in which 38% of pupils, 43% of teaching staff and 59% of non-teaching staff who participated in the investigation had anti-SARS-CoV-2 antibodies^27^. This study would suggest that high school aged children have similar susceptibility to SARS-CoV-2 infection as adults, and can transmit SARS-CoV-2 efficiently. The reasons why the observed onwards transmission from primary school aged children was lower than in adolescents warrant further investigation. Given that viral load among infected children and adults have been found to be similar^21,22^, milder symptoms among children may explain the reduced onward transmission. In households, we found a high prevalence of antibodies among parents and relatives of infected pupils (61% and 44%, respectively). Considering the low onward transmission from pupils in schools, the high prevalence figures observed in families more likely resulted from transmission from the parents to the child rather than the opposite, which has already been suggested by others^18^.

In adults, symptoms associated with SARS-CoV-2 infection were fever, cough, shortness of breath, ageusia, anosmia, headache, asthenia, muscle pain, sore throat, and diarrhea, all known features of the disease. Symptoms with highest predictive values for SARS-CoV-2 infection were anosmia and ageusia, as previously reported^32^. Symptoms were less specific in children, with only fatigue and diarrhea being associated with SARS-CoV-2 infection. Anosmia and ageusia were rarely seen (1% of children), and only in those aged 15 years or older. This study also gave an opportunity to estimate the proportion of asymptomatic forms among infected. Here, we found that the asymptomatic fraction was higher in children than in adults (41.4% vs 9.9%, respectively; P < 0.001).

Our results are limited by the short timeframe for studying the impact of the presence of infected pupils in schools before closure, which happened only two weeks after the first cases of COVID-19 developed in pupils. Still, the absence of well characterized viral spread in the primary school as opposed to what had been observed in the nearby high school at the same time suggests that children aged 6-11 years may be less contagious than high school aged pupils. Another limitation was the incomplete sampling of classes and families, preventing from a full exploration of viral circulation in the schools and households. The clinical findings of our investigation were also limited by the fact that information on symptoms was collected retrospectively, and that other respiratory viruses were circulating concurrently in the study population.

## Conclusion

The findings of our investigation are in line with other reports which suggest limited transmission of SARS-CoV-2 in primary schools. These findings suggest that reopening of primary schools can be considered carefully, with continuous monitoring of possible resurgence in infections and strategies to limit transmission such as hand hygiene, physical distancing, respiratory etiquette and masks for older children.

## Data Availability

All data generated or analysed during this study are included in this published article (and its supplementary information files)

## Acknowledgements

We would like to thank the Directorate General for Health for facilitating the initiation of the study, Santé publique France for linking with their investigation team, the Agence Régionale de Santé des Hauts de France, and the Academia of Amiens for their continuous support throughout the realization of the study, especially Ms. Delphine Maskara. We would like to thank the mayor of Crépy-en-Valois, Bruno Fortier and his team, Mrs Sandra Riveros, the local police, and local paramedical staff, Nathalie Bourgeois, Mélissa Mannevy, and Michaël Mannevy, for their help in the implementation of the study. We would like also to thank Nathalie de Parseval, Claire Dugast, Valentine Garaud, Soazic Gardais, Caroline Jannet, Fanny Momboisse, Isabelle Porteret, Hantaniaina Rafanoson, Sandrine Ropars who participated in the organization and the realization of the field investigation. Julian Buchrieser and Rémy Robinot who assisted in the development of serological tests. And finally Nathalie Jolly and Cassandre van Platen, who took care of the regulatory aspects of the study, and Christine Fanaud who informed the participants of their serological status.

